# Real-World Effectiveness of Maternal RSVpreF Vaccination and Durability of Infant Protection

**DOI:** 10.64898/2026.09.15.26363141

**Authors:** Hanmeng Xu, Camila Aparicio-Llorente, Aanchal Wats, Barbara L. Araujo, Julia Moniz Ganem, Isabela O. Oliva, Eugene D. Shapiro, Nina N. Brodsky, Paul L. Aronson, Nathan D. Grubaugh, Carrie L. Lucas, Linda M. Niccolai, Joshua L. Warren, Lee Kennedy-Shaffer, Virginia E. Pitzer, Daniel M. Weinberger, Carlos R. Oliveira

## Abstract

In this test-negative study, the effectiveness of the maternal RSVpreF vaccine among infants <6 months was 73.9% against medically attended respiratory syncytial virus (RSV) infection and 86.3% against RSV-associated hospitalization. Maternal RSVpreF and nirsevimab had comparable initial effectiveness, but effectiveness of RSVpreF waned approximately 1.7 times more rapidly.

## Introduction

Respiratory syncytial virus (RSV) is a leading cause of respiratory illness and hospitalization in infants [1]. Maternal RSVpreF vaccination was first licensed in the United States (U.S.) in 2023 after clinical trials demonstrated efficacy against severe RSV disease in infants through transplacental transfer of antibodies [2]. However, important questions remain regarding its effectiveness in routine clinical practice. Most postlicensure studies have focused on early infancy up to 6 months of age [3–6], leaving the durability of protection beyond the first few months incompletely characterized. In addition, evidence is limited on the effectiveness of maternal vaccines within current U.S. practice, where nirsevimab and other monoclonal antibodies are available as complementary protection.

To address these gaps, the primary aim of this study was to estimate the real-world effectiveness of maternal RSVpreF vaccination against medically attended RSV infection and RSV-associated hospitalization during the first 6 months of life. Secondary objectives were to characterize the durability of protection through 12 months of age and to examine patterns of waning alongside nirsevimab.

## Methods

### Data Sources and Study Population

We conducted a test-negative case-control study using linked electronic health records (EHRs) from mother–infant dyads within Yale New Haven Health System (YNHHS). Infants were included in the analysis if they were born between October 1, 2023, and September 30, 2025, and underwent RSV testing for an acute respiratory illness (ARI) during the first 12 months of life. To ensure that included infants were born to mothers eligible for RSVpreF vaccination, we further restricted the cohort to infants born at ≥32 weeks’ gestation and whose mothers were 32–36 weeks pregnant during the recommended September–January vaccination period.

We extracted from the EHR patient characteristics, encounter setting (inpatient/outpatient), and RSV test results through March 1, 2026. Maternal and infant immunization histories were ascertained from the EHR and Connecticut’s statewide immunization registry, to which Connecticut providers are required to report vaccinations. Only immunizations with a documented administration date were included. Detailed variable definitions are provided in the study dictionary (Table S1).

Cases were defined as infants with ARI who tested positive for RSV by polymerase chain reaction of nasopharyngeal samples, and controls as infants with ARI who tested negative for RSV. ARI was ascertained using free-text searches and structured EHR fields, including encounter diagnoses and problem lists. Multiple RSV tests from the same infant were consolidated into distinct illness episodes using a 14-day interval, as previously described [7].

### Statistical Analysis

For the primary analysis, we used multivariable logistic regression to estimate effectiveness against RSV infection among infants aged <6 months at the time of RSV testing. For analyses of effectiveness against RSV-associated hospitalization, cases and controls were restricted to infants with an inpatient admission within ±14 days of RSV testing.

Immunization status was modeled as a categorical variable representing maternal RSVpreF only, nirsevimab only, dual immunization, or neither product, with the latter serving as the reference. Maternal RSVpreF vaccination was counted if administered prior to delivery of the index pregnancy and nirsevimab if administered to the infant ≥7 days before RSV testing.

Multivariable models were adjusted for infant age at RSV testing, gestational age, calendar time, community RSV activity, and medical comorbidities. Community RSV activity was defined as the log-transformed weekly proportion of Connecticut emergency department visits attributed to RSV among children 1 to 4 years old, obtained from Epic Cosmos via the PopHIVE data platform. Effectiveness was calculated as (1 - odds ratio) * 100%.

The extent to which the protection from RSVpreF varied over time after birth was estimated using logistic regression within a Bayesian framework, similar to the model used in our previous studies [7]. Waning effectiveness of RSVpreF was compared to that of nirsevimab over time since immunization. A comprehensive description of the model structure is provided in the Supplementary Methods. We conducted several sensitivity analyses to evaluate the robustness of our findings to alternative analytic assumptions. These included testing a more restrictive maternal RSVpreF exposure definition and different assumptions about the structure of the component of the model that captures waning.

All data cleaning and analyses were conducted in R, version 4.3.1, with code documented in https://github.com/Hanmeng-Xu/RSV_3seasons_MV. The institutional review board (IRB) at the Yale School of Medicine approved the study (HIC:2000036550). This study follows the Strengthening the Reporting of Observational Studies in Epidemiology (STROBE) reporting guideline.

## Results

Between October 1, 2023 and March 1, 2026, a total of 10,489 medically attended ARI encounters among infants aged ≤12 months were identified at YNHHS (Figure S1). Of these, 3,466 were born in the YNHHS to mothers who were also eligible for the maternal RSVpreF vaccine during pregnancy and were included in our study. Among the eligible infants, 396 were cases testing positive for RSV, and 3,070 were controls. 1,682 (48.5%) infants received at least one of the two products, and 341 (9.8%) received both. Demographic and clinical characteristics are shown in Table S2. Median infant age at testing was 5.4 months, and 13.4% of episodes involved infants with at least one medical comorbidity.

### Effectiveness of RSVpreF and nirsevimab

Effectiveness of maternal RSVpreF vaccination alone against medically attended RSV infection in the first 6 months of life was 73.9% (95% confidence interval (CI), 32.8%, 92.3%), broadly consistent with estimates from prelicensure clinical trials and recent real-world studies [2–6,8–12] (Figure 1). Nirsevimab showed similar effectiveness against medically attended RSV infection at 67.9% (95% CI, 51.9%, 79.1%), and dual immunization had effectiveness of 46.8% (95% CI, −0.4%, 73.9%). For RSV-associated hospitalization, effectiveness was 86.3% (95% CI, 17.5%, 99.3%) for maternal RSVpreF vaccination, 89.5% (95% CI, 72.8%, 96.7%) for nirsevimab, and 89.4% (95% CI, 42.8%, 99.4%) for dual immunization (Figure S2A). Sensitivity analysis using a strict definition for RSVpreF vaccination showed similar results (Figure S3).

**Figure 1.**
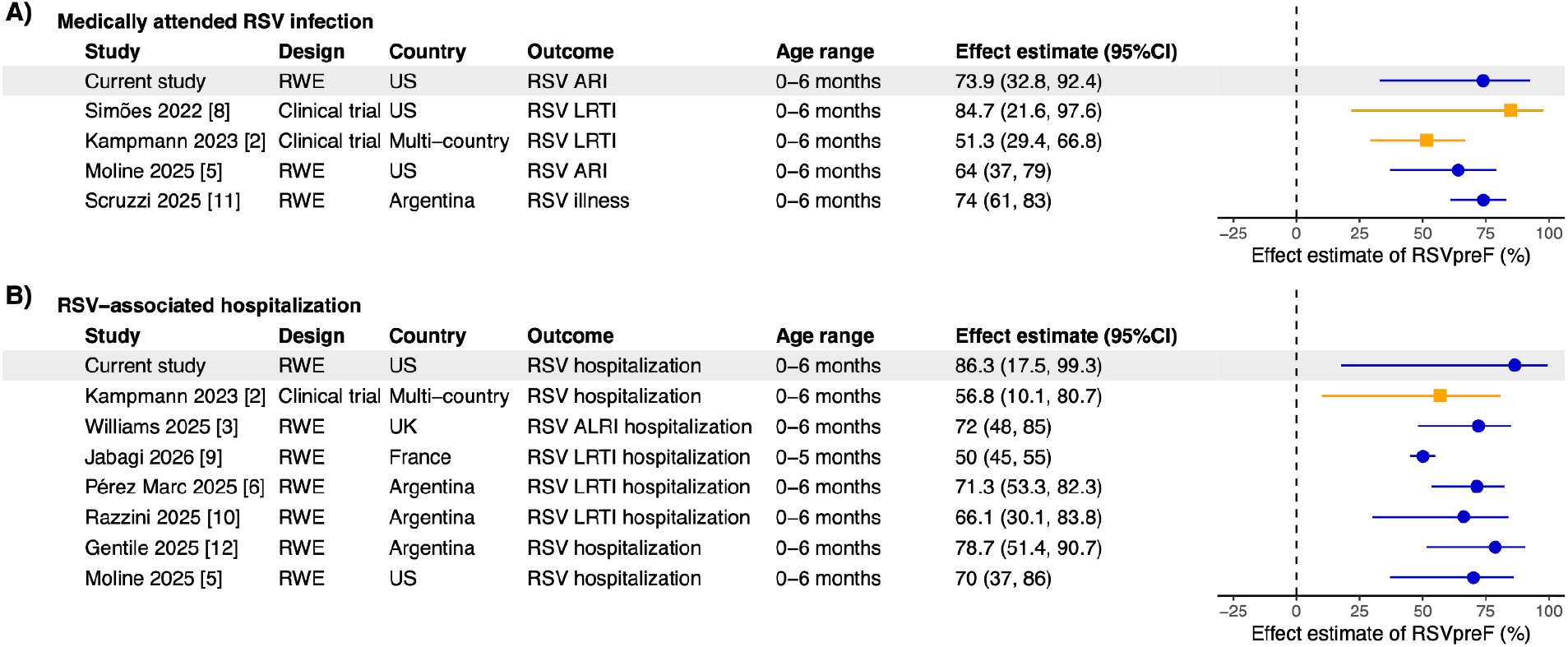
Protective effect of maternal RSVpreF vaccination alone across studies. Orange squares represent clinical trial efficacy and blue circles represent real-world effectiveness estimates; horizontal bars indicate 95% confidence intervals, and gray shading highlights the current study. ARI, acute respiratory illness; LRTI, lower respiratory tract infection; RSV, respiratory syncytial virus; RWE, real-world evidence.

### Waning effectiveness of maternal RSVpreF vaccine and nirsevimab

In the waning analysis, protection at 1 month was similar for maternal RSVpreF and nirsevimab, with effectiveness against RSV infection of 73.5% (95% credible interval (CrI), 39.0%, 92.3%) and 70.6% (95% CrI, 54.8%, 82.6%), respectively. By 6 months, modeled effectiveness had declined to 40.3% (95% CrI, −6.1%, 71.2%) for RSVpreF compared with 51.9% (95% CrI, 30.4%, 67.3%) for nirsevimab (Figure 2). Note that the structure imposed on the waning and priors used in the waning analysis means that the waning effectiveness estimates can be below those of the overall estimate in the previous section. The modeled decline in effectiveness was 6.2 (95% CrI, 2.3, 13.2) percentage points per month for maternal RSVpreF versus 3.6 (95% CrI, 1.4, 7.4) for nirsevimab, corresponding to a rate of waning approximately 1.7 (95% CrI, 0.5, 5.7) times faster with maternal vaccination. Findings were consistent across alternative Bayesian model structures (Figures S4).

**Figure 2.**
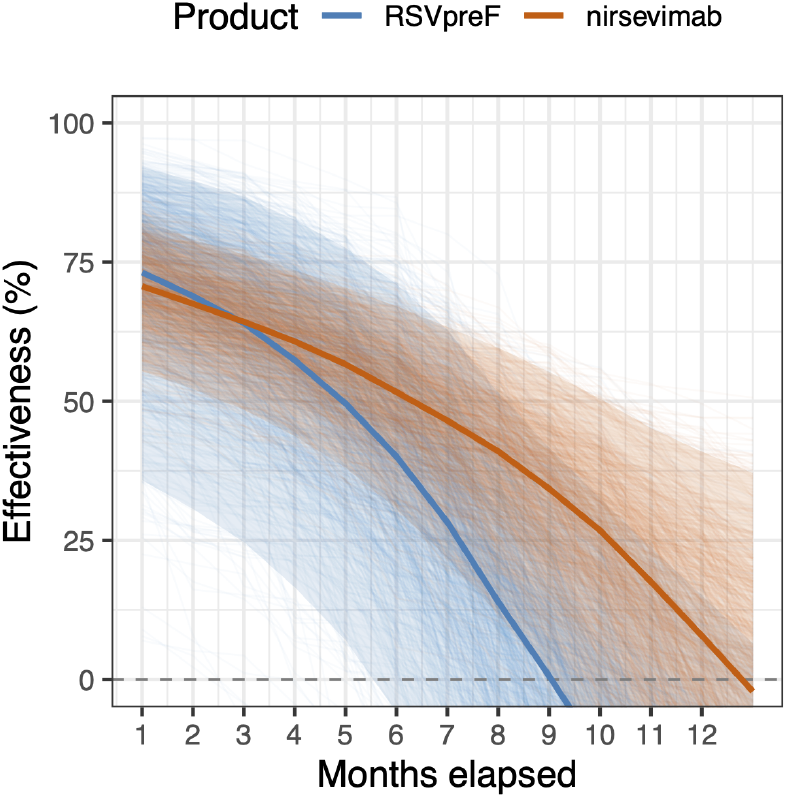
Modeled effectiveness against medically attended RSV infection over time. Thin lines show sampled posterior trajectories, solid lines show the posterior medians, and shaded areas indicate pointwise 95% credible intervals. The model included age at test as both a confounder and effect modifier of the immunization effect, and additionally adjusted for weekly RSV activity among 1 to 4 years old, gestational age at birth, and presence of at least one medical comorbidity.

## Discussion

In this test-negative case-control study spanning three RSV seasons, maternal RSVpreF vaccination was associated with a reduced risk of medically attended RSV infection and hospitalization during the first 6 months of life, consistent with prelicensure clinical trials [2,8]. Importantly, our analysis was designed as a pragmatic assessment of effectiveness under current U.S. practice, where maternal vaccination and infant immunization are used as complementary strategies. We found that effectiveness against hospitalization in early infancy was similar in magnitude across maternal vaccination, nirsevimab, and dual immunization, providing empirical evidence that the current U.S. approach to RSV prevention is protecting infants during their period of greatest vulnerability.

As evidence supporting maternal RSVpreF effectiveness accumulates, durability has become an increasingly important consideration, particularly given year-to-year variation in the timing of RSV circulation. Prior postlicensure studies have suggested declining effectiveness with increasing infant age but have had limited ability to characterize durability beyond the first few months of life or distinguish biologic waning from concurrent changes in RSV circulation [6,9,13]. By including infants tested through 12 months of age and accounting for these temporal factors in our models, we characterized waning of the maternal vaccine protection and examined this pattern alongside nirsevimab using a common analytic approach. Our data suggested divergence in the waning trajectories, with a steeper decline in effectiveness after maternal RSVpreF vaccination than after nirsevimab.

This pattern of waning is consistent with the known kinetics of maternally transferred RSV antibodies and nirsevimab. Prelicensure trials estimated a half-life of approximately 6 weeks for RSV-neutralizing antibodies transferred after maternal RSVpreF vaccination [14]. Thus, less than 10% of the antibody concentration present at birth would be expected to remain by 6 months. In contrast, nirsevimab was engineered with an Fc modification that enhances FcRn-mediated recycling, extending its half-life to approximately 10 weeks, about 1.7 times longer than that of maternally transferred antibodies [15]. This difference is directionally consistent with our estimates, which showed approximately 1.7-fold faster waning of effectiveness following maternal RSVpreF vaccination than nirsevimab. Further research is needed to determine whether the timing of maternal vaccination and infant immunization should be adapted to account for these differences in durability.

This study has limitations. First, the relatively small number of infants exposed to maternal RSVpreF, particularly among hospitalized infants and at older ages, limited the precision of some estimates and our ability to directly compare immunization strategies. Second, our exposure definition may limit direct comparability with studies that required vaccination at least 14 days before delivery. We chose this approach to provide a more pragmatic estimate of effectiveness and reflect real-world use of maternal immunization. Excluding or reclassifying pregnancies in which delivery limited transplacental antibody transfer may omit one of the principal ways the maternal strategy can fail in practice. Third, hospitalizations occurring outside YNHHS may not have been captured, resulting in potential outcome misclassification. Finally, receipt of maternal vaccination, nirsevimab, or both was not randomized and could have been influenced by factors not captured in our analysis. Comparisons between strategies should therefore be interpreted cautiously.

In conclusion, the current U.S. strategy of complementary maternal and infant immunization for RSV prevention was effective against early-life RSV infection and hospitalization. Protection waned over time, with point estimates suggesting a steeper decline following maternal immunization than nirsevimab, suggesting that durability may be important when determining how these strategies are implemented.

## Data Availability

External researchers can make written requests to the corresponding author for sharing of completely de-identified and aggregate-level data. Requests will be assessed on a case-by-case basis in consultation with the lead and co-investigators. All data sharing will abide by rules and policies defined by the involved parties. Data-sharing mechanisms will ensure that the rights and privacy of individuals participating in research will be protected at all times

