## Supplementary Materials for "Real-World Effectiveness of Maternal RSVpreF Vaccination and Durability of Infant Protection"

### Supplementary methods

#### Estimating the effectiveness of RSVpreF and nirsevimab by time elapsed

We evaluated the waning protection from RSVpreF over time since birth and waning protection from nirsevimab over time since immunization using logistic regression models within a Bayesian framework. The analytic population for the nirsevimab waning analysis comprised unimmunized infants and those who received nirsevimab alone. We further restricted immunized infants to infants who were immunized within the first month of life, so that nirsevimab conferred protection from approximately birth, comparable to the timing of protection conferred by maternal RSVpreF vaccination. The analytic population for maternal RSVpreF included those immunized and those "vaccinated" with RSVpreF.

##### Logistic regression model evaluating waning effectiveness

Waning effect was estimated by comparing those unimmunized to those immunized at the same age. We included age as both a confounder and a modifier in the logistic regression model.

For the $j$th test record in the analytic population, the observed case status (i.e., whether the patient tested positive or negative for RSV) followed a Bernoulli distribution, such that

$$Case\_Status_j \sim Bernoulli(p_j).$$

The age at RSV test of infants was categorized into monthly intervals. We created indicator variables to represent each time/age category: $age\_bin_{jn}$ $(n = 1, 2, 3..., 13)$ representing RSV test at 0 - <1 month, 1 - <2 months, … 11- < 12 months, and 12 months of age and above. For example, for an individual $j$ aged 20 days at RSV test (falls into 0-1 month), $age\_bin_{j1} = 1$ and $age\_bin_{jn} = 0$ for $n = 2, 3, ..., 13$. Immunization status was represented using a binary variable $vax$. The probability of an individual testing positive for RSV $p_j$, was modeled using a multivariable logistic regression framework as follows:

$$logit\left(p_j\right) = \sum_{n=1}^{N} \beta_n^a age\_bin_{jn} + vax_j * \sum_{n=1}^{N} \beta_n age\_bin_{jn} + \sum_{m=1}^{M} \alpha_m Z_{jm}$$

where $\beta_n$ is the immunization effect coefficient for each of the $N$ age bins and $\beta_n^a$ is the baseline effect for age. $Z_{jm}$ represents potential confounders, in which $m$ represents the number of confounders included in the regression model. We additionally included RSV activity (weekly RSV positivity rate among 1-4 years old), presence of at least one risk factor for severe RSV disease, and gestational age as confounders, and $\alpha_m$'$s$ represent the coefficients for the confounders.

Prior structures for immunization effect coefficients $\beta_n$

We tested waning models with different hierarchical structures for the effectiveness coefficients $\beta_n$, including the model with an imposed monotonic trend, model using basis spline (B-spline) structure, and autoregressive structure, described below.

**Model 1: Imposed a monotonic trend on $\beta_n$:**

Given the waning nature of passive immunity, we assume that the product effectiveness follows a non-increasing trend over time elapsed. This structure allowed relatively lower flexibility compared to the other models, allowing $\beta_n$ to either increase or remain unchanged over time, such that effectiveness could only decrease or plateau over time. To reflect this in the model, we imposed a monotonic structure on the regression coefficients $\beta_n$'s, such that

$$\beta_{n+1} = \beta_n + d_{n+1}, \; n = 1, 2, 3, \ldots$$

$$d_{n+1} \sim N^{+}(0, \tau_d^{-1})$$

$$\tau_d \sim Gamma(0.01, \; 0.01)$$

$d_{n+1}$ follows a truncated normal distribution, taking only non-negative values, and we used weakly informative priors on the increment $d_n$ over time. For the first coefficient $\beta_1$ (coefficient for the effectiveness for 0-1 month), we also used a weakly informative prior distribution:

$$\beta_1 \sim N(0, \; 100^2).$$

**Model 2: B-Spline model:**

In the B-spline model, the effectiveness coefficient $\beta_n$ was modeled as a linear combination of different “trends” over time since immunization:

$$\beta_n = \gamma_1 B_{n,1} + \gamma_2 B_{n,2} + \ldots. + \gamma_k B_{n,k}$$

in which $B$ is the basis spline matrix with a degree of freedom of $k$, $B_{n,i}$ is the $(n, i)th$ entry of $B$ (corresponding to time bin $n$ and column $k$), and $\gamma$'s are the spline coefficients. This ensured smoothness in the effectiveness estimates. An example of $B$ matrix with a degree of freedom of 6 is shown below:

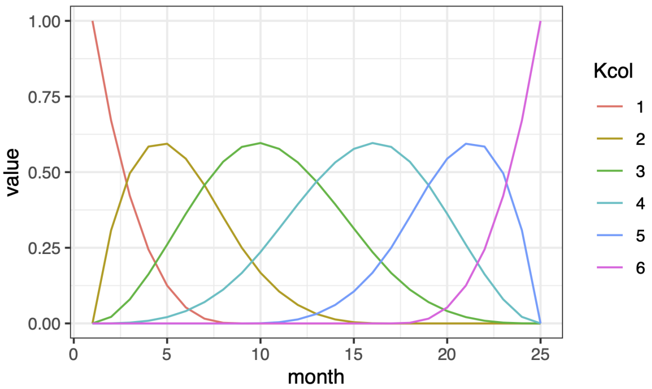


Each colored line ($B_k$) represents a trend with a specific shape over time and the estimated $\beta_n$ over time is a linear combination of these trends. We adopted a random walk structure on the spline coefficients $\gamma$'s:

$$\gamma_1 \sim N(0,\ 100^2)$$

$$\gamma_k \sim N(\gamma_{k-1},\ \tau_{spl}^{-1})$$

$$\tau_{spl} \sim Gamma(0.01,\ 0.01)$$

We tested models with the degree of freedom taking the values of 4,5, and 6, and compared the Deviance Information Criterion (DIC) across models. The difference in DIC across models was less than 2. We used the model with the degree of freedom of 6 in our main analysis.

**Model 3: Autoregressive $\beta_n$ with linear mean function:**

We also adopted a model structure without any imposed trend in the effectiveness over time, allowing the model to estimate the effectiveness coefficients with more flexibility. Instead, we added a first-order autoregressive structure (i.e., AR(1)) on the coefficients to prevent abrupt changes between neighboring coefficients and extended the AR(1) prior on the immunization effect over time by incorporating a linear mean function to stabilize the estimates, such that:

$$\beta_1 \sim N(0,\ [\tau_{arL}(1-\rho_{arL}^2)]^{-1})$$

$$\beta_n \sim N(\mu_0 + \mu_1(n-1) + \rho_{arL}\beta_{n-1},\ \tau_{arL}^{-1}),\ n = 2, 3, 4....$$

$$\mu_0, \mu_1 \sim N(0,\ 100^2)$$

$$\rho_{arL} \sim Uniform(-1,\ 1)$$

$$\tau_{arL} \sim Gamma(0.01,\ 0.01)$$

We separately evaluated the waning effectiveness of RSVpreF and nirsevimab using the above model and examined the effectiveness over time against medically attended RSV infection. Models were fitted via the rjags package in R version 4.3.1, in which we collected 5,000 samples from the posterior distribution after discarding the first 10,000 samples in the burn-in period. Convergence was evaluated using trace plots (Figure S5). The estimated effectiveness of RSVpreF and nirsevimab after a given period of time (for the time interval $n$) $IE_n$ was calculated as

$$IE_{mat,n} = \left(1 - e^{\beta_n^{mat}}\right) * 100\%,\ n = 1, 2, 3, \ldots$$
$$IE_{nir,n} = \left(1 - e^{\beta_n^{nir}}\right) * 100\%,\ n = 1, 2, 3, \ldots$$

Posterior medians and 95% quantile-based credible intervals were calculated from the collected posterior samples.

### Supplementary Figures

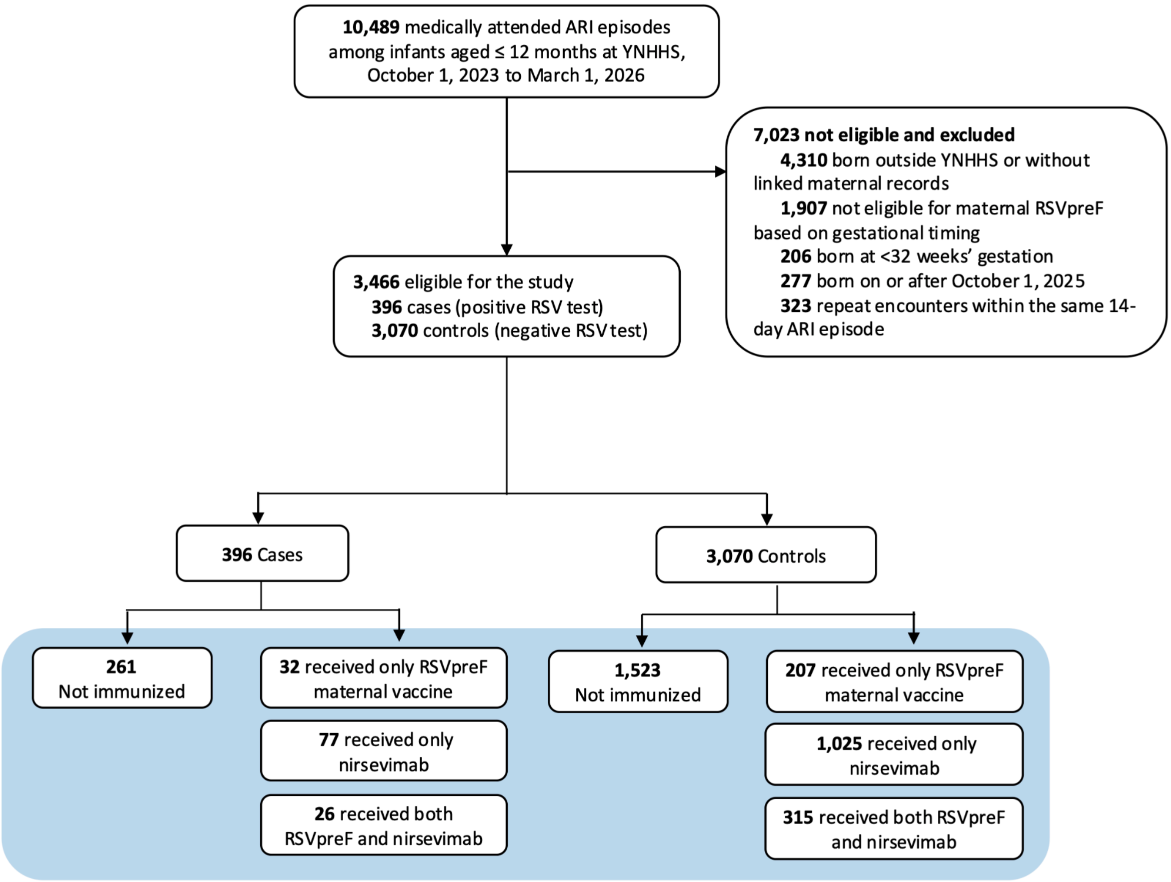


**Figure S1. Selection of cases and controls.**

**A) Effectiveness among infants aged < 6 months**

| Outcome prevented | Cases | Controls | Unadjusted Effectiveness | Adjusted Effectiveness |
|---|---|---|---|---|
| **Medically attended RSV infection** | | | | |
| Unimmunized | 168 | 880 | Reference | Reference |
| RSVpreF only | 4 | 103 | 79.7 (50.7, 93.8) | 73.9 (32.8, 92.3) |
| Nirsevimab only | 34 | 519 | 65.7 (50.2, 77) | 67.9 (51.9, 79.1) |
| Both RSVpreF and nirsevimab | 12 | 142 | 55.7 (21.6, 77.2) | 46.8 (−0.353, 73.[illegible] |
| **RSV−associated hospitalization** | | | | |
| Unimmunized | 68 | 201 | Reference | Reference |
| RSVpreF only | 1 | 23 | 87.1 (37.2, 99.3) | 86.3 (17.5, 99.3) |
| Nirsevimab only | 5 | 106 | 86.1 (67.6, 95.2) | 89.5 (72.8, 96.7) |
| Both RSVpreF and nirsevimab | 1 | 30 | 90.1 (52.6, 99.5) | 89.4 (42.8, 99.4) |

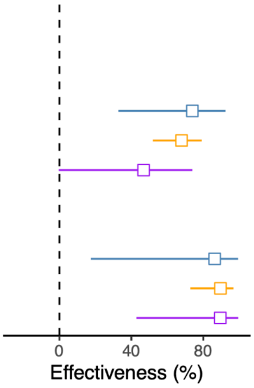


**B) Effectiveness among infants aged >= 6 months**

| Outcome prevented | Cases | Controls | Unadjusted Effectiveness | Adjusted Effectiveness |
|---|---|---|---|---|
| **Medically attended RSV infection** | | | | |
| Unimmunized | 93 | 643 | Reference | Reference |
| RSVpreF only | 28 | 104 | −86.1 (−195, −14.7) | −26.2 (−112, 26.3 |
| Nirsevimab only | 43 | 506 | 41.2 (14.6, 60.1) | 31.3 (−4.75, 55.4) |
| Both RSVpreF and nirsevimab | 14 | 173 | 44 (2.54, 70.1) | 36.9 (−19.7, 68.6) |
| **RSV−associated hospitalization** | | | | |
| Unimmunized | 14 | 59 | Reference | Reference |
| RSVpreF only | 4 | 7 | −141 (−819, 43.3) | 1.82 (−388, 81.8) |
| Nirsevimab only | 7 | 56 | 47.3 (−36.4, 81.2) | 38.9 (−131, 84.9) |
| Both RSVpreF and nirsevimab | 1 | 15 | 71.9 (−57.7, 98.5) | 81.2 (−113, 99.3) |

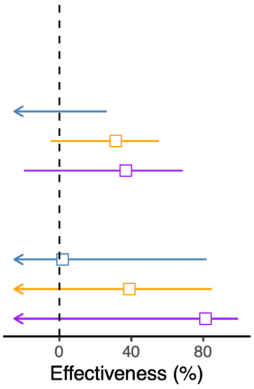


**C) Effectiveness among infants aged < 3 months**

| Outcome prevented | Cases | Controls | Unadjusted Effectiveness | Adjusted Effectiveness |
|---|---|---|---|---|
| **Medically attended RSV infection** | | | | |
| Unimmunized | 125 | 503 | Reference | Reference |
| RSVpreF only | 3 | 67 | 82 (50.5, 95.6) | 76.2 (30.5, 94.4) |
| Nirsevimab only | 26 | 281 | 62.8 (42.7, 76.6) | 68.6 (49.4, 81.1) |
| Both RSVpreF and nirsevimab | 7 | 80 | 64.8 (27, 85.5) | 64.3 (20.1, 86.1) |
| **RSV−associated hospitalization** | | | | |
| Unimmunized | 59 | 166 | Reference | Reference |
| RSVpreF only | 0 | 22 | NA | NA |
| Nirsevimab only | 5 | 68 | 79.3 (50.8, 93) | 86.6 (60.9, 96.3) |
| Both RSVpreF and nirsevimab | 1 | 25 | 88.7 (45, 99.4) | 93.6 (59.2, 99.7) |

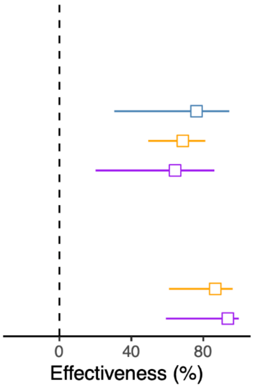


**Figure S2. Estimated effectiveness of different RSV immunization strategies among infants aged A) under 6 months, B) 6 to 12 months, and C) under 3 months.** The points and error bars represent medians and 95% confidence intervals of the estimates. Different immunization strategies were represented by different colors.

**A) Effectiveness among infants aged < 6 months**

| Outcome prevented | Cases | Controls | Unadjusted Effectiveness | Adjusted Effectiveness |
|---|---|---|---|---|
| **Medically attended RSV infection** | | | | |
| Unimmunized | 168 | 886 | Reference | Reference |
| RSVpreF only | 4 | 97 | 78.3 (47.2, 93.4) | 71.4 (25.9, 91.6) |
| Nirsevimab only | 35 | 543 | 66 (50.9, 77.1) | 67.8 (52, 78.9) |
| Both RSVpreF and nirsevimab | 11 | 118 | 50.8 (10.8, 75.5) | 39.4 (−17.2, 71.2) |
| **RSV−associated hospitalization** | | | | |
| Unimmunized | 68 | 204 | Reference | Reference |
| RSVpreF only | 1 | 20 | 85 (25.9, 99.2) | 84.7 (5.27, 99.2) |
| Nirsevimab only | 5 | 113 | 86.7 (69.2, 95.4) | 89.6 (73, 96.7) |
| Both RSVpreF and nirsevimab | 1 | 23 | 87 (36.3, 99.3) | 87.6 (32, 99.3) |

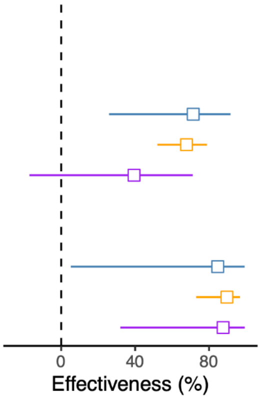


**B) Effectiveness among infants aged >= 6 months**

| Outcome prevented | Cases | Controls | Unadjusted Effectiveness | Adjusted Effectiveness |
|---|---|---|---|---|
| **Medically attended RSV infection** | | | | |
| Unimmunized | 93 | 644 | Reference | Reference |
| RSVpreF only | 28 | 103 | −88.2 (−198, −16) | −26.2 (−112, 26.3 |
| Nirsevimab only | 45 | 530 | 41.2 (15, 59.9) | 30.6 (−5.43, 54.8) |
| Both RSVpreF and nirsevimab | 12 | 149 | 44.2 (−0.558, 71.6) | 39.8 (−18.5, 71.4) |
| **RSV−associated hospitalization** | | | | |
| Unimmunized | 14 | 60 | Reference | Reference |
| RSVpreF only | 4 | 6 | −186 (−1040, 34.2) | −20.9 (−533, 78.2 |
| Nirsevimab only | 7 | 60 | 50 (−29.1, 82.1) | 49.7 (−92.6, 88.1) |
| Both RSVpreF and nirsevimab | 1 | 11 | 61 (−127, 98) | 6.72 (−1350, 96.9 |

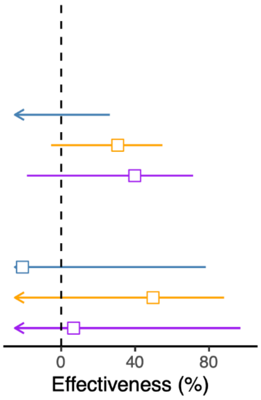


**C) Effectiveness among infants aged < 3 months**

| Outcome prevented | Cases | Controls | Unadjusted Effectiveness | Adjusted Effectiveness |
|---|---|---|---|---|
| **Medically attended RSV infection** | | | | |
| Unimmunized | 125 | 509 | Reference | Reference |
| RSVpreF only | 3 | 61 | 80 (44.8, 95.2) | 73.7 (22.6, 93.9) |
| Nirsevimab only | 26 | 297 | 64.4 (45.2, 77.6) | 69.6 (51.1, 81.7) |
| Both RSVpreF and nirsevimab | 7 | 64 | 55.5 (6.77, 81.8) | 53.1 (−6.41, 81.9) |
| **RSV−associated hospitalization** | | | | |
| Unimmunized | 59 | 169 | Reference | Reference |
| RSVpreF only | 0 | 19 | NA | NA |
| Nirsevimab only | 5 | 72 | 80.1 (52.8, 93.3) | 86.9 (61.6, 96.4) |
| Both RSVpreF and nirsevimab | 1 | 21 | 86.4 (32.6, 99.2) | 92.5 (52, 99.6) |

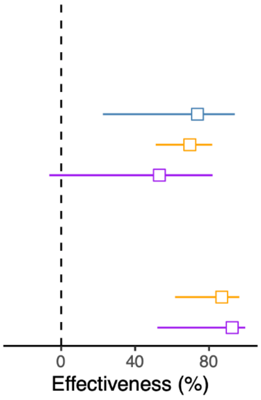


**Figure S3. Estimated effectiveness of different RSV immunization strategies among infants aged A) under 6 months, B) 6 to 12 months, and C) under 3 months, using a strict definition for RSVpreF vaccination.** In this sensitivity analysis, we defined RSVpreF vaccination as receiving RSVpreF at least 14 days before the delivery of the infant. The points and error bars represent medians and 95% confidence intervals of the estimates. Different immunization strategies were represented by different colors.

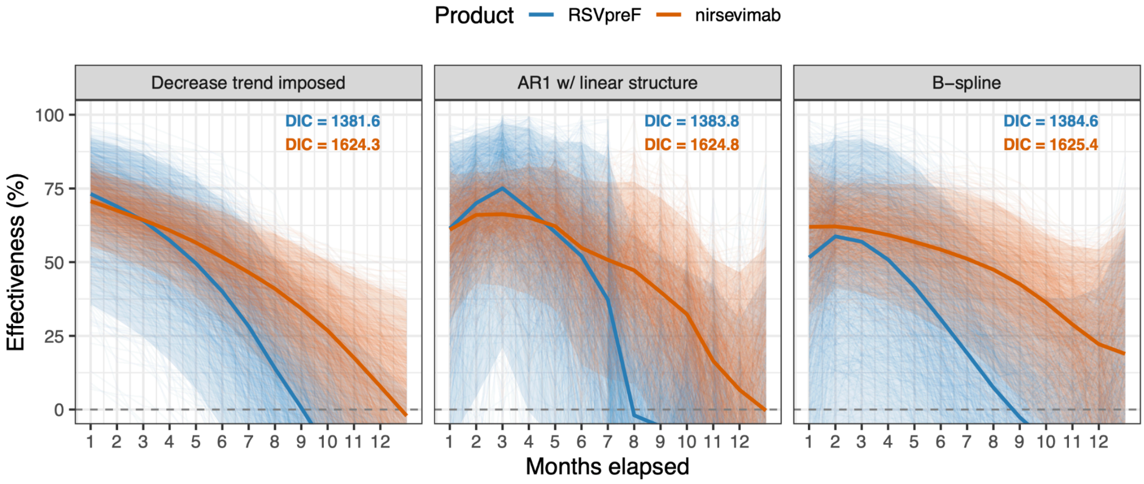


**Figure S4. Estimated effectiveness of RSVpreF and nirsevimab over time in preventing medically attended RSV infection, using alternative prior structures for immunization effect coefficients.** We randomly sampled 1,000 samples from the posterior of each model; each line represents one sampled trajectory. The shaded areas represent 95% credible intervals, and the lines represent the posterior medians of effectiveness over time (for RSVpreF: time since birth; for nirsevimab: time since immunization). The three panels present estimates from three different model structures, as described in the Supplementary Methods. Deviance Information Criterion (DIC) values are labeled.

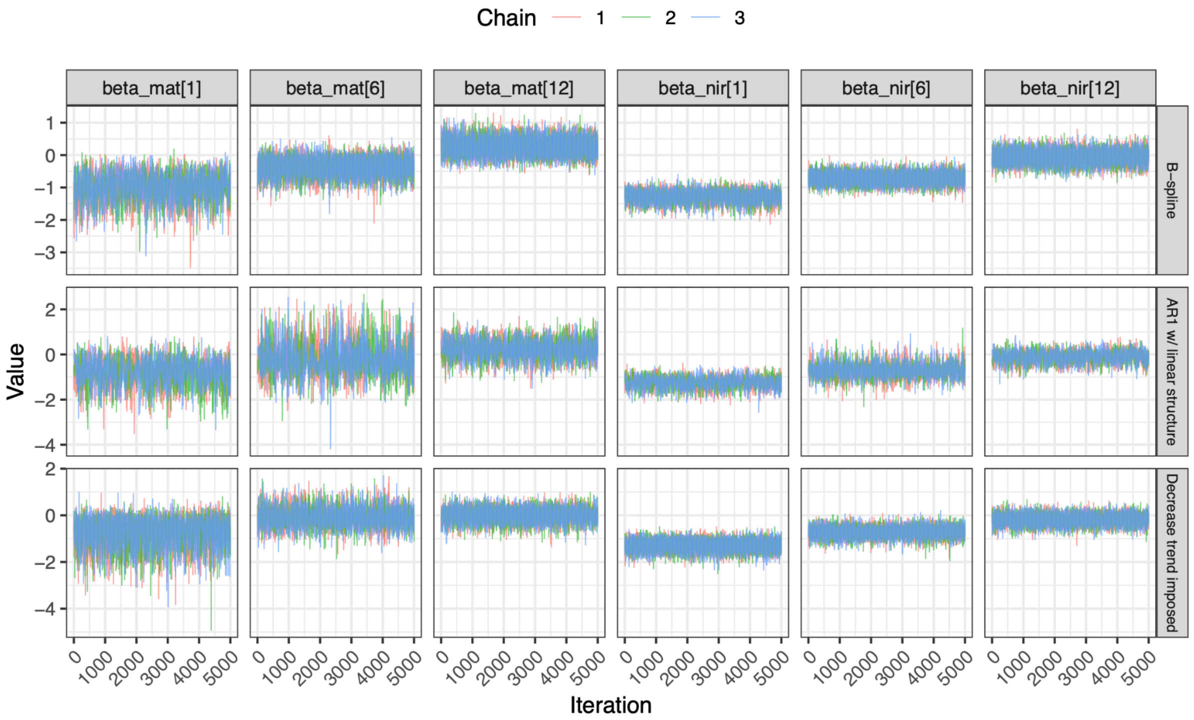


**Figure S5. Trace plots of the estimated coefficient of the maternal RSVpreF vaccine for each time interval since birth.** beta_mat[1], beta_mat[6], beta_mat[12] represent the maternal RSVpreF effectiveness coefficient against medically attended RSV infection 1, 6, 12 months after birth, and beta_nir[1], beta_nir[6], beta_nir[12] represent the nirsevimab effectiveness coefficient.

### Supplementary Tables

#### Table S1. Free-text patterns and ICD-10-CM codes used to identify risk factors for severe RSV diseases and acute respiratory illness (ARI).

| Category | Target item | Identification codes and free-text patterns |
|---|---|---|
| Medical comorbidities | Asthma | Asthma, reactive airway disease, J45 |
| | Congenital heart disease | Congenital heart disease, coarctation of aorta, congenital heart, D-TGA, double outlet right ventricle, dextrocardia, DORV, cardiac abnormality, ventricular septa, mitral regurgitation, coarctation of aorta, hypoplastic aortic arch, atrial septal defect, heart abnormality, cardiomegaly, patent ductus arteriosus, tetralogy of fallot, atrioventricular canal (AVC), persistent left superior vena cava, aortic arch anomaly, wolff-Parkinson-White (WPW) syndrome, supraventricular tachycardia, partial AV canal, atrial septa, supraventricular tachycardia, partial anomalous pulmonary venous return, aortic coarctation, congenital coronary artery fistula to pulmonary artery, peripheral pulmonic stenosis, tricuspid atresia with normal great arteries, Q20, Q21, Q22, Q23, Q24, Q25 |
| | Immunodeficiency | Liver transplanted, transplant, leukemia, D80, D81, D82, D83, D84, Z94, B20, B21, B22, B23, B24, C91, C92, C93, C94, C95 |
| | Down syndrome | Down syndrome, trisomy 21, Q90 |
| | Chronic lung disease of prematurity | Bronchopulmonary dysplasia, P27.1, P27.0, P27.8, P27.9 |
| Acute respiratory illness (ARI) | / | Fever, febrile; Cough; Wheezing; Abnormal breathing, breathing difficulty, difficulty breathing, shortness of breath, breathing problem, dyspnea, apnea, tachypnea; Abnormal breathing, breathing difficulty, difficulty breathing, breathing problem, shortness of breath, cough, acute obstructive laryngitis, pharyngitis, otitis media, upper respiratory infection, upper respiratory tract infection, upper resp. tract infection, upper resp. infection, croup, nasopharyngitis, pain in throat, reactive airway disease, nasal congestion, sore throat, URI, laryngeal stridor; Bronchiolitis, bronchitis, laryngotracheobronchitis, bronchospasm, acute chest syndrome, acute hypoxic respiratory failure, pneumonia, hypoxia, hypoxemia, lower resp. tract infection, lower resp. infection, lower respiratory tract infection, lower respiratory infection, hypoxemic respiratory failure, acute respiratory failure with hypoxia.<br>R50; R05; R06.2; R06; J00, J01, J02, J03, J04, J05, J06; J12, J13, J14, J15, J16, J17, J18, J20, J21, J22 |

**Table S2. Characteristics of infants aged <12 months at acute respiratory illness encounters, by RSV infection status**

| Characteristic | Overall, No. (%) | RSV-Positive, N = 396 | RSV-Negative, N = 3,070[1] | SMD[2] |
|---|---|---|---|---|
| Sex | | | | 0.05 |
| Female | 1,455 (42.0%) | 157 (39.6%) | 1,298 (42.3%) | |
| Male | 2,011 (58.0%) | 239 (60.4%) | 1,772 (57.7%) | |
| Race and ethnicity | | | | 0.10 |
| Black, non-Hispanic | 662 (19.1%) | 65 (16.4%) | 597 (19.4%) | |
| White, non-Hispanic | 822 (23.7%) | 105 (26.5%) | 717 (23.4%) | |
| Other race, non-Hispanic[4] | 215 (6.2%) | 24 (6.1%) | 191 (6.2%) | |
| Hispanic or Latino | 1,701 (49.1%) | 194 (49.0%) | 1,507 (49.1%) | |
| Unknown | 66 (1.9%) | 8 (2.0%) | 58 (1.9%) | |
| Age at RSV test (months) | | | | 0.00 |
| Mean (SD) | 5.8 (3.7) | 5.8 (4.0) | 5.8 (3.6) | |
| Median (IQR) | 5.4 (2.3, 9.3) | 4.4 (2.1, 9.8) | 5.5 (2.4, 9.2) | |
| Season of test | | | | 0.21 |
| 2023/24 | 944 (27.2%) | 126 (31.8%) | 818 (26.6%) | |
| 2024/25 | 1,733 (50.0%) | 208 (52.5%) | 1,525 (49.7%) | |
| 2025/26 | 789 (22.8%) | 62 (15.7%) | 727 (23.7%) | |
| Month tested | | | | 0.95 |
| Dec-Jan | 1,060 (30.6%) | 241 (60.9%) | 819 (26.7%) | |
| Feb-Mar | 661 (19.1%) | 52 (13.1%) | 609 (19.8%) | |
| Oct-Nov | 602 (17.4%) | 79 (19.9%) | 523 (17.0%) | |
| Off-season months | 1,143 (33.0%) | 24 (6.1%) | 1,119 (36.4%) | |
| Gestational age at birth | | | | 0.08 |
| Median (IQR) | 39.0 (37.0, 39.0) | 39.0 (38.0, 39.0) | 39.0 (37.0, 39.0) | |
| Birth weight (g) | | | | 0.22 |
| Median (IQR) | 3,228 (2,869, 3,588) | 3,298 (3,026, 3,658) | 3,218 (2,849, 3,578) | |
| Insurance status | | | | 0.09 |
| Private | 927 (26.7%) | 119 (30.1%) | 808 (26.3%) | |
| Public | 2,536 (73.2%) | 277 (69.9%) | 2,259 (73.6%) | |
| Unknown | 3 (0.1%) | 0 (0.0%) | 3 (0.1%) | |
| History of prematurity (<37 wk) | 453 (13.1%) | 36 (9.1%) | 417 (13.6%) | -0.14 |
| Cardiovascular Condition | 243 (7.0%) | 15 (3.8%) | 228 (7.4%) | -0.16 |
| Comorbid condition[5] | 464 (13.4%) | 44 (11.1%) | 420 (13.7%) | -0.08 |
| Hospitalized | 598 (17.3%) | 101 (25.5%) | 497 (16.2%) | 0.23 |
| RSV immunization | | | | 0.38 |
| Unimmunized | 1,784 (51.5%) | 261 (65.9%) | 1,523 (49.6%) | |
| RSVpreF only | 239 (6.9%) | 32 (8.1%) | 207 (6.7%) | |
| Nirsevimab only | 1,102 (31.8%) | 77 (19.4%) | 1,025 (33.4%) | |
| Both RSVpreF and nirsevimab | 341 (9.8%) | 26 (6.6%) | 315 (10.3%) | |

[1]Values are No. (%) unless otherwise indicated in the Characteristic column.
[2]Standardized Mean Difference: the difference in means between case and control participants in units of the pooled SD. Covariates with an absolute standardized mean difference greater than 0.2 were considered to have important imbalances.
[3]Self-reported race and ethnicity.
[4]Including American Indian or Native American, Asian, Middle Eastern or Northern African, and Pacific Islander by self-reporting.
[5]Having at least 1 of the following conditions recorded in the infant's medical history or diagnosis records: (1) asthma, (2) immunodeficiency (e.g., transplantation history or leukemia), (3) cardiac diseases (including congenital heart diseases diagnosed at birth or any reporting of heart conditions), (4) pulmonary diseases, (5) Down syndrome. Observations represent distinct ARI episodes; an infant could contribute more than one episode.

### Table S3. Characteristics of tested infants receiving different combinations of RSV prophylactic products.

| Characteristic | **Overall**, N = 3,466[1] | **Unimmunized**, N = 1,784[1] | **RSVpreF only**, N = 239[1] | **Nirsevimab only**, N = 1,102[1] | **Both RSVpreF and nirsevimab**, N = 341[1] |
|---|---|---|---|---|---|
| **Sex** | | | | | |
| Female | 1,455 (42.0%) | 753 (42.2%) | 86 (36.0%) | 480 (43.6%) | 136 (39.9%) |
| Male | 2,011 (58.0%) | 1,031 (57.8%) | 153 (64.0%) | 622 (56.4%) | 205 (60.1%) |
| **Race and ethnicity[2]** | | | | | |
| Black, non-Hispanic | 662 (19.1%) | 311 (17.4%) | 41 (17.2%) | 252 (22.9%) | 58 (17.0%) |
| White, non-Hispanic | 822 (23.7%) | 449 (25.2%) | 91 (38.1%) | 203 (18.4%) | 79 (23.2%) |
| Other race, non-Hispanic[3] | 215 (6.2%) | 105 (5.9%) | 13 (5.4%) | 78 (7.1%) | 19 (5.6%) |
| Hispanic or Latino | 1,701 (49.1%) | 877 (49.2%) | 89 (37.2%) | 550 (49.9%) | 185 (54.3%) |
| Unknown | 66 (1.9%) | 42 (2.4%) | 5 (2.1%) | 19 (1.7%) | 0 (0.0%) |
| **Age at RSV test (months)** | | | | | |
| Mean (SD) | 5.8 (3.7) | 5.4 (3.6) | 6.3 (3.9) | 6.1 (3.7) | 6.5 (3.8) |
| Median (IQR) | 5.4 (2.3, 9.3) | 4.9 (2.1, 8.6) | 7.1 (2.3, 9.8) | 6.0 (2.7, 9.6) | 6.8 (3.0, 10.1) |
| **Season of test** | | | | | |
| 2023/24 | 944 (27.2%) | 679 (38.1%) | 55 (23.0%) | 176 (16.0%) | 34 (10.0%) |
| 2024/25 | 1,733 (50.0%) | 815 (45.7%) | 143 (59.8%) | 591 (53.6%) | 184 (54.0%) |
| 2025/26 | 789 (22.8%) | 290 (16.3%) | 41 (17.2%) | 335 (30.4%) | 123 (36.1%) |
| **Month tested** | | | | | |
| Dec-Jan | 1,060 (30.6%) | 535 (30.0%) | 78 (32.6%) | 340 (30.9%) | 107 (31.4%) |
| Feb-Mar | 661 (19.1%) | 305 (17.1%) | 43 (18.0%) | 257 (23.3%) | 56 (16.4%) |
| Oct-Nov | 602 (17.4%) | 339 (19.0%) | 37 (15.5%) | 170 (15.4%) | 56 (16.4%) |
| Off-season months | 1,143 (33.0%) | 605 (33.9%) | 81 (33.9%) | 335 (30.4%) | 122 (35.8%) |
| **Gestational age at birth** | | | | | |
| Mean (SD) | 38.3 (1.7) | 38.4 (1.6) | 38.6 (1.2) | 38.0 (1.9) | 38.2 (1.6) |
| Median (IQR) | 39.0 (37.0, 39.0) | 39.0 (37.0, 39.0) | 39.0 (38.0, 39.0) | 39.0 (37.0, 39.0) | 39.0 (37.0, 39.0) |
| **Birth weight (g)** | | | | | |
| Median (IQR) | 3,228 (2,869, 3,588) | 3,248 (2,928, 3,591) | 3,348 (3,033, 3,748) | 3,158 (2,739, 3,518) | 3,248 (2,828, 3,588) |
| **Insurance status** | | | | | |
| Private | 927 (26.7%) | 509 (28.5%) | 117 (49.0%) | 208 (18.9%) | 93 (27.3%) |
| Public | 2,536 (73.2%) | 1,274 (71.4%) | 122 (51.0%) | 893 (81.0%) | 247 (72.4%) |
| Unknown | 3 (0.1%) | 1 (0.1%) | 0 (0.0%) | 1 (0.1%) | 1 (0.3%) |
| **Hospitalized** | 598 (17.3%) | 342 (19.2%) | 35 (14.6%) | 174 (15.8%) | 47 (13.8%) |
| **Preterm** | 453 (13.1%) | 194 (10.9%) | 14 (5.9%) | 198 (18.0%) | 47 (13.8%) |
| **Cardiac diseases** | 243 (7.0%) | 109 (6.1%) | 15 (6.3%) | 91 (8.3%) | 28 (8.2%) |
| **Medical comorbidities[4]** | 464 (13.4%) | 250 (14.0%) | 32 (13.4%) | 144 (13.1%) | 38 (11.1%) |

[1]Values are No. (%) unless otherwise indicated in the Characteristic column.
[2]Self-reported race and ethnicity.
[3]Including American Indian or Native American, Asian, Middle Eastern or Northern African, and Pacific Islander by self-reporting.
[4]Have at least 1 of the following conditions recorded in the infant's medical history or diagnosis records: (1) asthma, (2) immunodeficiency (e.g., transplantation history or leukemia), (3) cardiac diseases (including congenital heart diseases diagnosed at birth or any reporting of heart conditions), (4) pulmonary diseases, (5) Down syndrome.
